# HealthCall: Smartphone Enhancement of Brief Interventions to Reduce Heavy Drinking In HIV Care

**DOI:** 10.1101/2020.11.25.20234328

**Authors:** Deborah Hasin, Efrat Aharonovich, Barry Zingman, Malka Stohl, Claire Walsh, Jennifer C. Elliott, David Fink, Justin Knox, Sean Durant, Raquel Menchaca, Anjali Sharma

## Abstract

**Background:** Heavy drinking among people living with HIV (PLWH) worsens their health outcomes and disrupts their continuum of care. Brief interventions to reduce heavy drinking in primary care are effective, but in heavy-drinking PLWH, more extensive intervention may be needed. Lengthy interventions are not feasible in most HIV primary care settings, and patients seldom follow referrals to outside treatment. Utilizing visual and video features of smartphone technology, we developed the “HealthCall” app to provide continued engagement after brief intervention, in order to reduce drinking and improve other aspects of HIV care while making minimal demands on providers.

**Methods:** Alcohol-dependent patients at a large urban HIV clinic were randomized to one of three groups: (1) Motivational Interviewing (MI) plus HealthCall (n=39), (2) NIAAA Clinician’s Guide (CG) plus HealthCall (n=38), or (3) CG-only (n=37). Baseline drinking-reduction interventions were ∼25 minutes, with brief (10-15 min) check-in sessions at 30 and 60 days. HealthCall involved daily use of the smartphone for 3-5 min/day, covering drinking and other aspects of the prior 24 hours. Outcomes assessed at 30 and 60 days, and 3, 6 and 12 months, included drinks per drinking day, drinks per day, and days drank, using the Timeline Followback. Analysis were conducted using generalized linear mixed models with pre-planned contrasts.

**Results:** Study retention was excellent (85%-94% across timepoints) and unrelated to treatment arm or patient characteristics. During treatment, patients in MI+HealthCall drank less than others (p=0.07-0.003). However, at 6 and 12 months, drinking was lowest among patients who had been in CG+HealthCall (p=0.04-0.06).

**Conclusion:** During treatment, patients in MI+HealthCall drank less than patients in the CG conditions. However, at 6 and 12 months, drinking was lower among patients in CG+HealthCall. Given the importance of drinking reduction and the low costs and time required for HealthCall, pairing HealthCall with brief interventions within HIV clinics merits widespread consideration.

## Introduction

Heavy alcohol use is associated with alcohol use disorder (AUD), as well as substantial morbidity and mortality^1,2^. For the 1.2 million people living with HIV (PLWH) in the United States, AUDs have similar or higher prevalence that of the general population^3,4^, with high levels of problem drinking in HIV primary care^5,6^. Heavy drinking among PLWH can worsen heart disease, exacerbate comorbid infections such as tuberculosis and Hepatitis C, and may accelerate cognitive decline^7^. Alcohol also negatively affects the HIV continuum of care, including engagement in care, treatment, and viral suppression^8,9^. In addition, HIV providers appear to spend less time with heavy drinkers and have poorer communication with them^10^. Beyond direct risks to patients, alcohol is associated with high-risk sex,^11^ which increases risk of HIV transmission^12^. Thus, effective interventions to decrease heavy drinking among PLWH are critical to improving their health and to HIV prevention. Because heavy-drinking medical patients seldom follow referrals to outside addiction specialists^13,14^, brief, effective interventions that can be administered within the HIV medical setting are needed.

Brief drinking-reduction interventions are effective at decreasing drinking in general primary care settings^15^, although alcohol dependent patients often require more extensive intervention^14,16,17^. Despite the importance of decreasing heavy drinking in PLWH, a growing literature of trials assessing alcohol interventions among PLWH shows that many interventions either lack efficacy or are very time-consuming^18,19^. Some HIV alcohol primary care interventions have been designed for easier dissemination, e.g., those based on the Screening and Brief Intervention with Referral to Treatment (SBIRT) model^20-22^ and interventions utilizing technology^23,24^. Interventions facilitated by technology are especially timely in the era of COVID-19, where telehealth has been used in HIV clinics to limit risk of COVID-19 infection^25^. In order to provide extended intervention with little added demand on provider time, we developed the HealthCall application, which leverages the visual and video features of smartphone technology to promote brief, continued patient engagement between sessions (see Figure 1)^16,17,26^. After a brief baseline intervention interview, patients engage with the HealthCall app for self-monitoring to make daily self-reports of drinking and other health behaviors for 60 days, and receive immediate positive reinforcement (“Good job!”). HealthCall utilizes the self-report data to provide users with personalized feedback and additional positive reinforcement at 30- and 60-days in a brief feedback session with a clinic provider.

**Figure 1.**
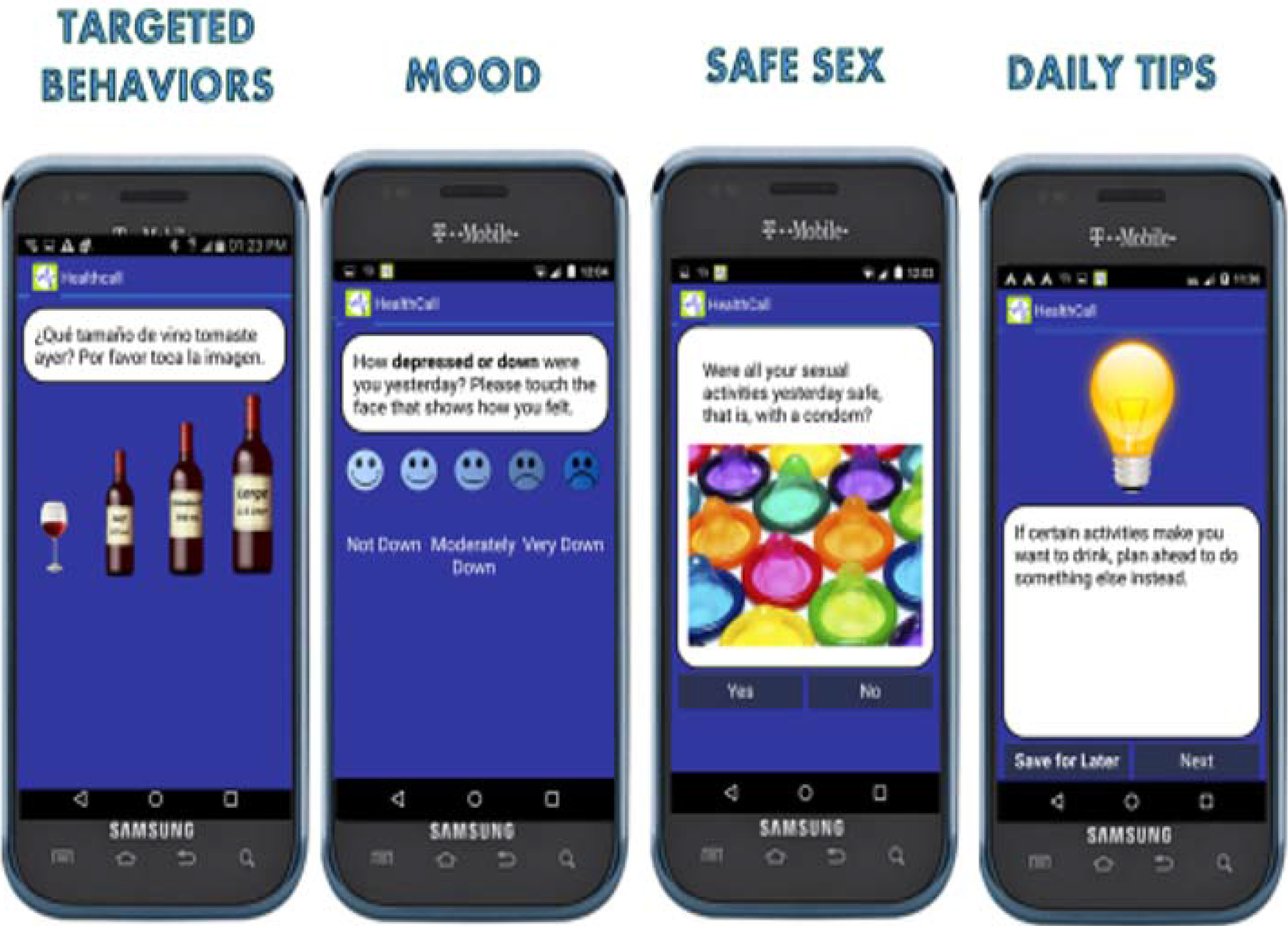
Examples of HealthCall Interactive Screens.

In earlier studies, HealthCall administered via Interactive Voice Response (IVR) technology was shown to successfully enhance the efficacy of brief Motivational Interviewing (MI), an evidence-based drinking-reduction intervention to motivate behavior change^27-35^. MI is broadly effective in helping patients change drinking and other risk behaviors in medical settings^36^. In our earlier study, MI-only and MI+HealthCall were compared to a health education-only control among 258 urban, HIV primary care patients who screened positive for heavy drinking, 48.4% of whom were alcohol dependent.^16^ In the alcohol dependent patients,^16^ MI+HealthCall was superior to MI-only and control conditions for drinking reduction at end-of-treatment (60 days). At 6- and 12-month follow-ups, alcohol dependent patients in MI+HealthCall continued to report less drinking than those in the other arms^16^. We have since migrated HealthCall technology to the smartphone^37^, which provides numerous advantages with features that can be added to improve patient engagement in the intervention^37^.

While MI is effectively enhanced by HealthCall, implementing MI has challenges. MI requires lengthy training (1-3 days);^38^ careful supervision;^39,40^ and skill, which is strongly related to its outcome.^41^ Time constraints and limited MI proficiency among staff without a counseling background are barriers to its implementation.^38^ We were therefore interested in testing whether HealthCall could enhance a less demanding brief intervention, making the intervention more scalable and facilitating dissemination. The evidence-based Clinician’s Guide (CG),^13,42,43^ designed and recommended by the National Institute on Alcoholism and Alcohol Abuse (NIAAA) for use by medical personnel,^42^ including HIV settings,^44^ is such an intervention. CG requires little training and staff time.^45^ It includes a flow-chart set of recommended^46^ brief drinking-reduction techniques whose efficacy over non-treatment conditions in non-dependent patients is well documented.^27,47-53^

In order to test HealthCall as an enhancer of efficacy when paired with CG or MI, we conducted a 1:1:1 randomized trial targeting drinking reduction among alcohol dependent patients receiving care in an urban, HIV primary care clinic. Arm 1 received CG-only, Arm 2 received CG and HealthCall, and Arm 3 received MI and HealthCall. Outcomes examined drinking reduction using quantity metrics: drinks per drinking day (primary outcome) and drinks per day. While the interventions were all oriented towards harm-reduction rather than abstinence, we also explored days drank, as days abstinent or drinking is a common outcome in alcohol clinical trials literature^54^, and non-drinking days allow the liver to heal^55^.

## Method

### Participants

Participants were recruited from and enrolled at a large urban infectious disease primary care clinic in New York City. Inclusion criteria included confirmed HIV infection, age≥18 years old, meeting DSM-IV criteria for current alcohol dependence^56^, drinking >4 drinks on at least one occasion in the past 30 days, and English- or Spanish speaking. Exclusion criteria included previous participation in a HealthCall study, active psychosis, active violent, homicidal, or suicidal thoughts, active alcohol withdrawal, gross cognitive impairment, hearing or vision impairment that precluded smartphone use, and plans to leave New York during the study period. Participants provided written informed consent. Institutional review boards at the New York State Psychiatric Institute and Montefiore Hospital, Bronx, New York approved all procedures.

### Procedures

Potentially eligible patients were informed about the study by IRB approved flyers in the clinic waiting room and/or by their providers, who referred them to meet with a study coordinator for written informed consent and assessment of eligibility. Of 180 individuals screened in-person with the Psychiatric Research Interview for Substance and Mental Disorders for DSM-IV alcohol and drug dependence (PRISM-IV-Computerized Version) (Hasin et al., 2006), 114 met eligibility, completed baseline assessments, and were randomized (see Study CONSORT Figure 2). In a parallel three-arm 1:1:1 randomized design, participants were assigned to one of three conditions: CG-only (n=37), CG+HealthCall (n=38), or MI+HealthCall (n=39). Randomization was stratified on drug use severity, depression, and unstable housing using urn randomization^57^. All baseline assessments were completed prior to randomization. After each study visit, participants were compensated via ClinCards for their completed assessments. All participants also received a study smartphone or the equivalent value in gift cards at their 60-day visit.

**Figure 2.**
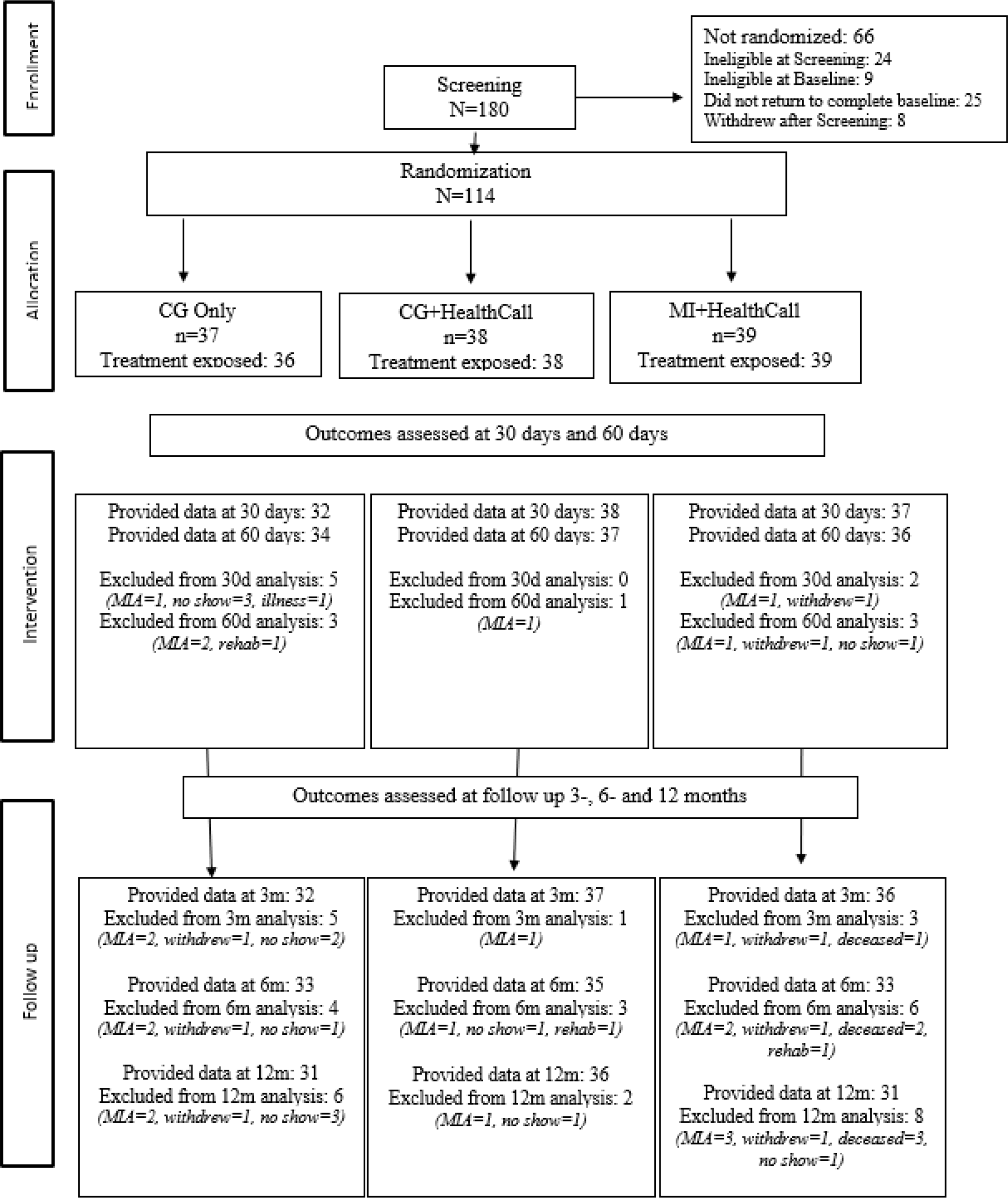
Study CONSORT

### Interventions

Study counselors, part of the regular clinic staff, delivered the interventions after receiving training in their delivery. A clinic health educator delivered the MI. An RN or a physician-fellow delivered the CG. The baseline MI and CG sessions took equivalent amounts of time (∼25 minutes). While delivering CG in the CG-only and CG+HealthCall arms, the counselors were blinded to participants’ assignment to HealthCall; about 20 minutes into the CG session, a study coordinator texted the counselor to inform them if the patient was randomly assigned to receive HealthCall. If assigned, the counselor then introduced the HealthCall app to the patient. This two-step randomization process kept counselors blinded to the HealthCall assignment until after the CG was delivered to avoid knowledge of the assignment influencing the way counselors delivered CG. Intervention sessions were audiotaped and reviewed for quality assurance. Participants returned at 30 and 60 days after the baseline intervention for assessments and a brief booster session (10-15 minutes). Total duration of the treatment period was 60 days across all three arms. Further face-to-face follow-up assessments were conducted at 3, 6, and 12 months after baseline.

### Clinician’s Guide (CG)

The evidence-based CG^13,42,43^ was designed and recommended by the National Institute on Alcoholism and Alcohol Abuse (NIAAA) for use by providers such as medical personnel,^33^ including those in HIV care settings.^44^ Consisting of a flow-chart to guide a set of recommended drinking-reduction techniques^46^, CG requires little training and staff time.^45^ The CG was adapted for the study to address slightly lower drinking reduction goals due to HIV infection, and also to address HIV medication adherence. At 30- and 60-day visits, participants met with the clinic staff member again for a booster session to review drinking and to reassess goals. In the CG-only arm, discussion of drinking in the prior 30 days was based on patients’ recall. In the CG+HealthCall arm, discussion was based on a graphed report of daily drinking data generated from the HealthCall app.

### Motivational interviewing (MI)

The brief MI session followed standard MI techniques^58^, e.g., exploring ambivalence about drinking reduction, health consequences of drinking, and setting a drinking goal for the next 30 days. At the 30- and 60-day visits, in a brief booster session, drinking behaviors during the previous 30 days were discussed based on graphed report of daily drinking data generated from the HealthCall app. Participant’s drinking reduction goals were reviewed, and new goals set if patients wished to do so.

### HealthCall application

Participants randomized to use HealthCall received an Android smartphone on which the HealthCall application and a pre-paid calling and data plan were installed. Study counselors introduced participants to the HealthCall application by explaining its purpose, instructing them in its use, and allowing them to try it on-site. Patients were asked to use HealthCall daily for the next 60 days. HealthCall is proposed to work through two features: self-monitoring and personalized feedback. ***Self-Monitoring***: Patients were asked to use HealthCall application daily (∼3-5 min each day). The HealthCall script includes self-monitoring questions in English or Spanish (participant’s preference) about the previous 24 hours^37^. The interactive queries asked about ‘yesterday’ (morning, afternoon, evening) to ensure consistent reporting periods regardless of the time participants accessed HealthCall. HealthCall queried drinking patterns (beverage types; number and size of drinks for each type), reasons for drinking or abstaining that day, as well as medication adherence, drug use, mood, and stress. ***Personalized feedback*:** Patient’s daily HealthCall data were automatically transmitted to a secure server, compiled, and used to produce personalized graphs representing the number of standard drinks patients drank each reported day relative to their drinking-reduction goal. The 30 and 60-day booster sessions included a review and discussion of these graphs. At the end of the 30-day booster visit, any revised drinking goals were entered into the app by interventionist. Participants were then asked to continue using HealthCall for another 30 days. At the end of the 60-day session (end of treatment), options for continued alcohol care were discussed, and referrals offered if requested.

### Measures

At baseline, 30 and 60 days during treatment, and at 3, 6 and 12 months after treatment, participants completed an interviewer-administered computer-assisted personal interview (CAPI). Assessments took ∼60 min administered by a trained independent study assessor. The CAPI assessed demographic characteristics, DSM-IV substance and psychiatric disorders, and other medical and HIV-related health variables. To assess detoxification needs for participants with physical dependence, alcohol withdrawal was assessed at baseline, and at 30- and 60-days visits with the Clinical Institute Withdrawal Assessment of Alcohol revised (CIWA-Ar)^59^. Alcohol use was assessed at all visits with a 30-day timeline-follow-back (TLFB)^60^. Study counselors were blind to the TLFB assessment results, and participants were informed of this. To calculate standard drinks, participants were shown the NIAAA “What’s a Standard Drink” conversion chart, with drink types and glass sizes^61^. If assessments were missed, during the next visit, a retrospective TLFB was administered to cover missed time periods. This retrospective method captures data with good reliability^62,63^. Self-reports of alcohol use were validated with breathalyzers.

### Outcomes

Our drinking outcomes reflected the harm-reduction rather than abstinence focus of the interventions. The primary outcome was mean number of drinks per drinking day in the prior 30 days (NumDD), selected due to the potential for liver toxicity/damage from large drinking quantities on days that patients drank^55,64^. We also analyzed mean drinks per day (Drinks/Day), which combines amounts consumed on drinking days with zero scores on days patients did not drink. Also included was days drank (DaysD), since days drinking versus abstinent is a common outcome in alcohol clinical trials. These outcomes were assessed using the TLFB (regarding the past 30 days) for all time-points.

### Statistical analysis

Baseline characteristics (e.g., demographic characteristics and alcohol use prior to baseline) were examined descriptively among the whole sample and by treatment group. Differences in characteristics by treatment group were tested using non-parametric Wilcoxon tests for continuous variables and Chi-square tests for categorical variables.

Change in alcohol use (NumDD, Drinks/day and DaysD) were analyzed using TLFB data from assessments during treatment (30 and 60 days) and from the follow-up assessments (3, 6, and 12 months). Generalized linear mixed models (GLMM) were used to model the effects of time, treatment, and the interaction of time x treatment on the outcomes using SAS PROC GLIMMIX^65^. Models controlled for baseline level of alcohol use and used a lognormal distribution to model the outcomes. Pre-planned contrasts were used to compare treatment groups (CG+HealthCall vs. CG-only; MI+HealthCall vs. CG-only; MI+HealthCall vs. CG+HealthCall). All tests were two-sided with p<0.05 indicating significance, also noting trends toward significance at p<0.10. Participants were all analyzed in their originally assigned treatment group. Incident Rate Ratios (IRR) and associated confidence intervals were used to represent effect sizes since they are suitable for non-normally distributed variables. IRRs reflect the ratio of expected count (e.g., number of drinks), or rate, of the outcome in one treatment group compared to the reference group at each timepoint.

## Results

### Sample characteristics

Of the 114 enrolled participants, 58% were male, 75% were African American, 28% were Hispanic, and 62% had less than a high school education (See Table 1). The mean age was 47.5 years (SD = 10 years) and the mean number of years of HIV infection was 18.6 (SD =7.6). Thirty-seven participants were randomized to CG-Only, thirty-eight to CG+HealthCall, and thirty-nine to MI+HealthCall. One patient randomized to CG-Only did not keep an appointment for the baseline intervention and thus was not exposed to any protocol intervention; this participant was not included in the analyses. None of the baseline demographic characteristics differed significantly by treatment condition.

**Table 1:** Demographics by treatment condition.

|  |  | All<br>(n=114) | Clinician<br>Guide Only<br>(n=37) | Clinician<br>Guide +<br>HealthCall<br>(n=38) | MI +<br>HealthCall<br>(n=39) |  |
| --- | --- | --- | --- | --- | --- | --- |
|  |  | % (n) | % (n) | % (n) | % (n) | p-value <sup>a</sup> |
| Sex |  |  |  |  |  |  |
|  | Male | 57.9 (66) | 51.4 (19) | 65.8 (25) | 56.4 (22) | 0.44 |
|  | Female | 42.1 (48) | 48.6 (18) | 34.2 (13) | 43.6 (17) |  |
| Ethnicity |  |  |  |  |  |  |
|  | Hispanic | 28.1 (32) | 32.4 (12) | 26.3 (10) | 25.6 (10) | 0.77 |
|  | Non-Hispanic | 71.9 (82) | 67.6 (25) | 73.7 (28) | 74.4 (29) |  |
| Race |  |  |  |  |  |  |
|  | Black | 75.4 (86) | 73.0 (27) | 79.0 (30) | 74.4 (29) | 0.82 |
|  | Non-Black | 24.6 (28) | 27.0 (10) | 21.1 (8) | 25.6 (10) |  |
| Education |  |  |  |  |  |  |
|  | Less than High<br>School | 62.3 (71) | 67.6 (25) | 50.0 (19) | 69.2 (27) | 0.16 |
|  | High school grad/<br>GED or higher | 37.7 (43) | 32.4 (12) | 50.0 (19) | 30.8 (12) |  |
|  |  | M (SD) | M (SD) | M (SD) | M (SD) |  |
| Age |  |  |  |  |  |  |
|  |  | 47.5 (10.0) | 46.5 (10.1) | 46.7 (10.7) | 49.1 (9.3) | 0.37 |
| Years of HIV<br>infection |  |  |  |  |  |  |
|  |  | 18.6 (SD =7.6) | 20.3 (7.3) | 16.3 (7.9) | 19.1 (7.2) | 0.07 |

<sup>a</sup> P-values: Chi-square for categorical variables and Kuskall-Wallis test for continuous variables.

At baseline, participants drank on average 13.9 (SD=8.2) of the previous 30 days (DaysD). Their average number of drinks consumed on drinking days was 7.7 (SD=4.5) (NumDD). Their average drinks per day (Drinks/Day) was 3.5 (SD=3.0). There were no differences between treatment conditions on these three drinking variables at baseline, ps=0.36-0.55 (Table 2).

**Table 2:**
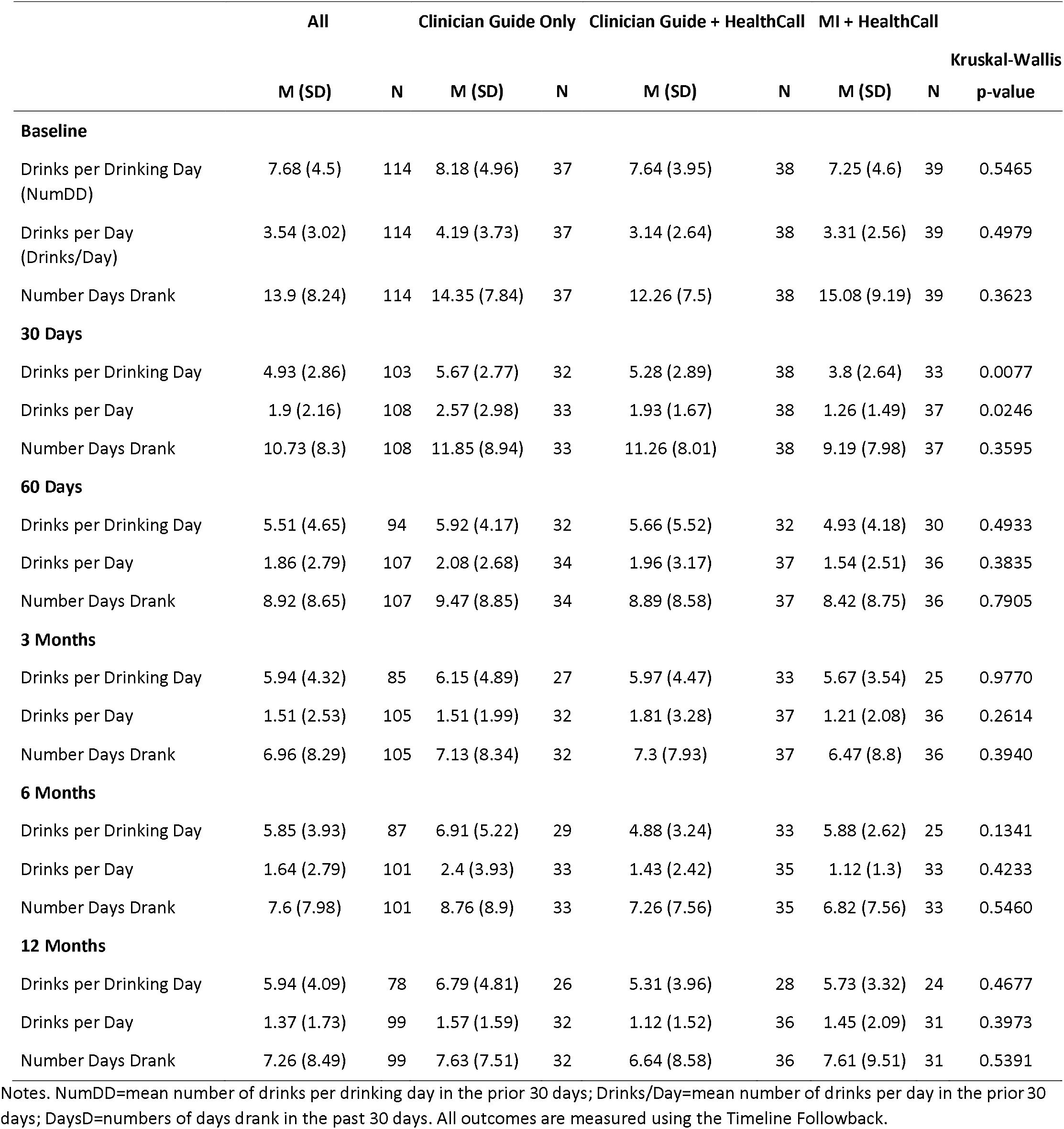
Alcohol outcomes from Timeline Follow-back, including Number of drinks per drinking day (NumDD), Drinks/Day, and Days Drank (DaysD).

### Retention and HealthCall Engagement

Overall, study retention was high across the treatment conditions. Of the 114 participants, 98 (86%) completed the study (n=31 in CG-Only, n=36 in CG+HealthCall, and n=31 in MI+HealthCall). Retention did not differ significantly by treatment condition or any baseline characteristic.

Participants reported high utilization of HealthCall. Among patients in MI+HealthCall, the median HealthCall call rate was 89.17%. Among participants in CG+HealthCall, the median HealthCall call rate was 87.50%.

### Drinking outcomes

#### Mean drinks per drinking day (NumDD)

Overall, NumDD decreased from baseline (see Figure 3a). Across conditions, NumDD was 7.7 (SD=4.5) at baseline, 4.9 (SD=2.9) at 30 days and 5.5 (SD=4.7) at 60 days (end-of-treatment), and then remained about 5.9 at 3 months, 6 months, and 12 months. Between-group contrasts are shown in Table 3. At 30 days, reduction in NumDD was 38% greater among patients in MI+HealthCall than among patients in CG-only (IRR=0.62, p=0.003) and 36% greater than among patients in CG+HealthCall (IRR=0.64, p=0.004). At 60 days, the difference between MI+HealthCall versus CG-only was at a trend level of significance (IRR=0.75, p=0.07). Although groups looked similar at 3 months, by 6 months, CG+HealthCall’s reduction in NumDD was 25% greater than among patients in CG-only (IRR=0.75, p=0.07), and at 12 months, patients in CG+HealthCall had a 29% greater reduction in NumDD than patients in the CG-only, a finding that was statistically significant (IRR=0.71, p=0.04).

**Table 3:** Treatment effects on alcohol outcomes at 30 days, 60 days, 3, 6, and 12 months using a Generalized Linear Mixed Model (GLMM).

|  |  | Contrasts |  |  |  |  |  |
| --- | --- | --- | --- | --- | --- | --- | --- |
|  |  | CG+HealthCall vs.<br>CG Only |  | MI+HealthCall<br>vs. CG Only |  | MI+HealthCall<br>vs. CG+HealthCall |  |
|  |  | *IRR* | *p* | *IRR* | *p* | *IRR* | *p* |
| **Drinks per drinking day** |  |  |  |  |  |  |  |
|  | 30 days | 0.96 | 0.81 | 0.62 | 0.003 | 0.64 | 0.004 |
|  | 60 days | 0.95 | 0.73 | 0.75 | 0.07 | 0.79 | 0.14 |
|  | 3 months | 1.00 | 0.97 | 0.97 | 0.86 | 0.96 | 0.82 |
|  | 6 months | 0.75 | 0.066 | 0.94 | 0.72 | 1.26 | 0.16 |
|  | 12 months | 0.71 | 0.0355 | 0.81 | 0.23 | 1.15 | 0.40 |
| **Drinks per day** |  |  |  |  |  |  |  |
|  | 30 days | 1.11 | 0.71 | 0.58 | 0.06 | 0.52 | 0.02 |
|  | 60 days | 1.07 | 0.83 | 0.83 | 0.54 | 0.78 | 0.40 |
|  | 3 months | 0.94 | 0.84 | 0.83 | 0.54 | 0.88 | 0.66 |
|  | 6 months | 0.55 | 0.04 | 0.73 | 0.30 | 1.33 | 0.35 |
|  | 12 months | 0.50 | 0.02 | 0.64 | 0.15 | 1.28 | 0.42 |
| **Number of days drank** |  |  |  |  |  |  |  |
|  | 30 days | 1.09 | 0.68 | 0.81 | 0.31 | 0.75 | 0.15 |
|  | 60 days | 1.12 | 0.59 | 0.98 | 0.91 | 0.87 | 0.52 |
|  | 3 months | 0.94 | 0.76 | 0.75 | 0.20 | 0.80 | 0.30 |
|  | 6 months | 0.73 | 0.13 | 0.69 | 0.09 | 0.94 | 0.78 |
|  | 12 months | 0.71 | 0.11 | 0.70 | 0.11 | 0.98 | 0.94 |

*Note.* Each model is adjusted for the baseline level of the outcome variable and uses a log link. IRR= Incidence risk ratio.

**Figure 3a:**
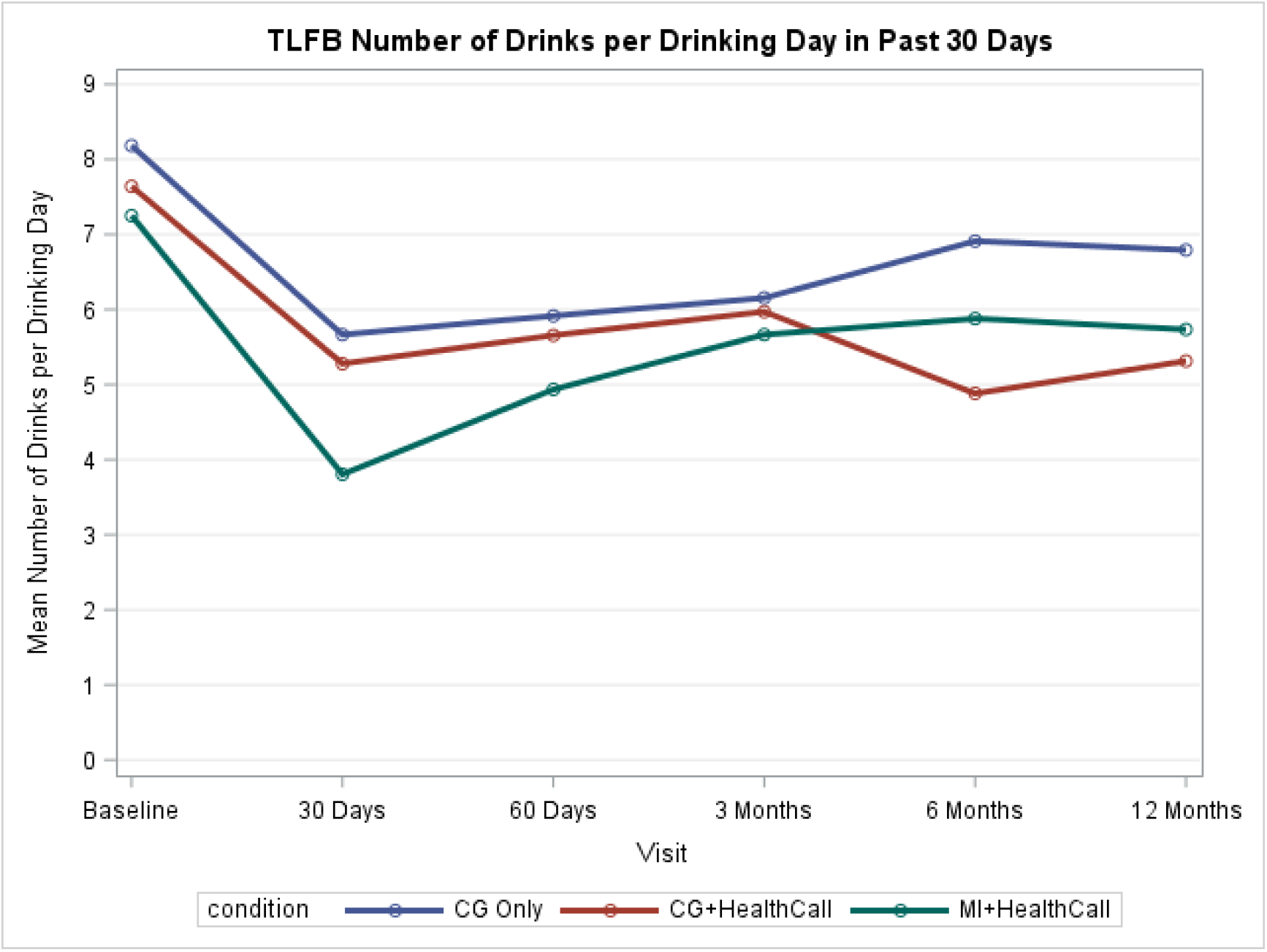
TLFB Number of Drinks per Drinking Day in Past 30 Days (NumDD)

**Figure 3b:**
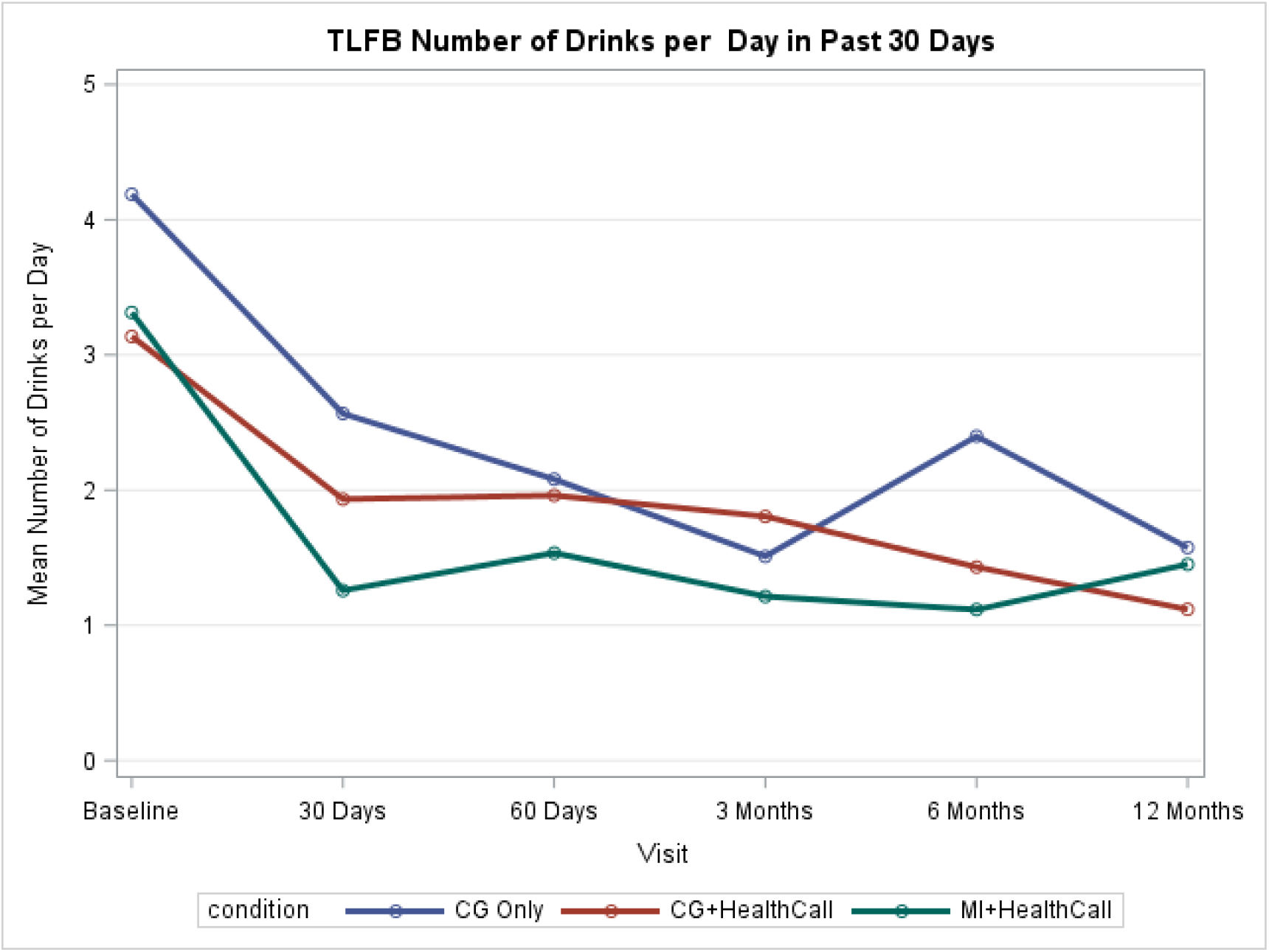
TLFB Drinks per day (Drinks/day) in Past 30 Days

**Figure 3c:**
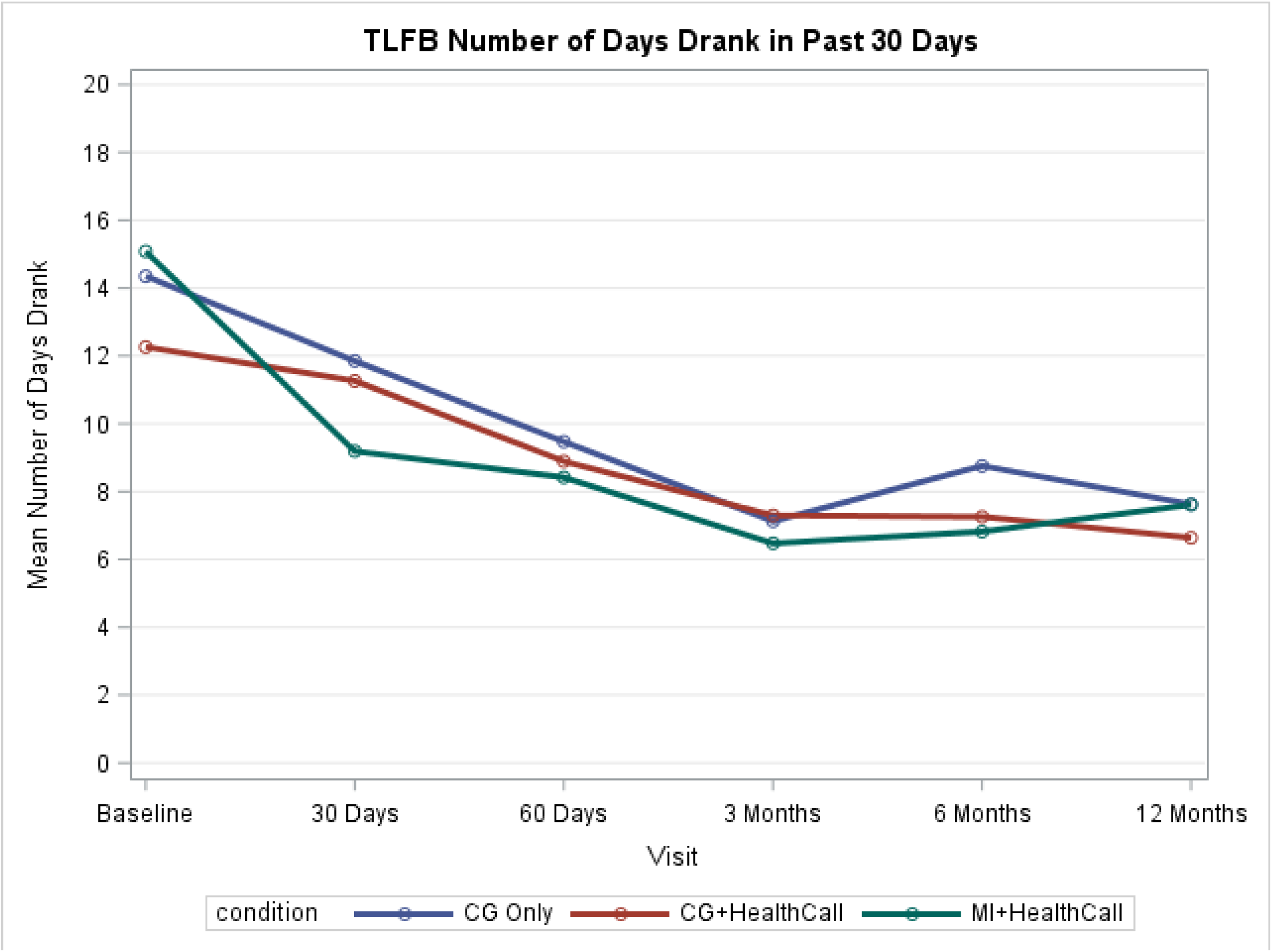
TLFB Number of Days Drank (DaysD) in Past 30 Days

#### Drinks per day (Drinks/day)

Overall, drinks/day decreased during the study (see Figure 3b). Across conditions, drinks/day was 3.5 (SD =3.0) at baseline, dropped to approximately 1.9 at 30 and 60 days (end-of-treatment), and then dropped to the 1.4-1.6 range at 3 months, 6 months, and 12 months. MI+HealthCall evidenced the lowest drinks/day values at all follow-up periods except 12 months; CG-only evidenced the highest drinks/day values at all timepoints except 3 months. At 30 days, reduction in drinks/day was 48% greater among patients in MI+HealthCall than among patients in CG+HealthCall (IRR=0.52, p=0.02) and 42% greater among MI+HealthCall than among patients in CG-Only (IRR=0.58, p=0.06). Differences were not significant at 60 days or 3 months, but by 6 months, CG+HealthCall evidenced a 45% greater reduction than CG-only (IRR=0.55, p=0.04), and by 12 months, this was a 50% greater reduction (IRR=0.50, p=0.02).

#### Number of days drank (DaysD)

Overall, DaysD decreased during the study (see Figure 3c). Across conditions, DaysD was 13.9 (SD =8.2) at baseline, dropped to 10.7 (SD=8.3) at 30 days and 8.9 (SD=8.7) at 60 days (end-of-treatment), and then dropped to the 7.0-7.6 range at 3 months, 6 months, and 12 months. Although there were no significant differences by treatment condition, MI+HealthCall maintained the lowest DaysD from 30 days until 6 months, and the difference between MI+HealthCall and CG-Only was marginal at this 6-month time point (IRR=0.69, p=0.09).

## Discussion

Reducing heavy drinking among PLWH is an important clinical and public health concern. Effective interventions that do not require extensive staff time or specialized training offer the best possibilities for dissemination. Accordingly, this randomized controlled trial evaluated the efficacy of HealthCall as a technologically—based enhancement to either a brief (25 min) Motivational Interview or session of advice/motivation guided by the NIAAA Clinician’s Guide, compared with use of the Clinician’s Guide only, in reducing drinking among alcohol dependent PLWH. During treatment, patients in MI+HealthCall drank less than patients in either the CG+HealthCall or the CG-Only arm. However, at 6 and 12 months, drinking was lowest among patients who had received CG+HealthCall.

Among alcohol dependent patients receiving care in an HIV primary care clinic, a brief-intervention strategy plus HealthCall was associated with drinking reductions compared to CG only. In earlier work, we successfully utilized an IVR version of HealthCall^66^. The emergence of smartphone technology facilitated migrating HealthCall to a more user-friendly and technologically advanced platform that offered greater capacity for patient engagement and positive reinforcement^37^. This smartphone version of HealthCall was the version tested in the present study. The results supports the use of HealthCall for treatment enhancement that compares favorably with more personnel- and resource-heavy interventions previously developed to reduce drinking among PLWH^18^. The current version of HealthCall also addresses medication adherence and potential triggers for drinking (e.g., mood, drug use, reasons for drinking, to be reported in future papers), potentially encouraging PLWH to consider drinking in relation to their health as a whole. Preliminary findings for HealthCall suggest that brief intervention plus HealthCall also helps improve HIV medication adherence among PLWH^67^.

Study limitations are noted. Recruitment was limited to one large clinic in an urban setting, potentially limiting generalizability to other geographic regions such as rural areas, where patients might benefit from an intervention that can be used without extensive travel. Inclusion criteria required DSM-IV alcohol dependence, limiting generalizability to non-dependent drinkers or those who may meet the lower threshold criteria for DSM-5 alcohol use disorder. Further work in larger or multi-site samples could more thoroughly address HealthCall results in other groups not well represented here. In addition, patients with severe alcohol withdrawal were excluded because medically-assisted treatment for withdrawal is warranted for such cases. Thus, the study does not provide information about patients initially presenting with severe withdrawal, who might benefit from HealthCall once their withdrawal has been treated medically and resolved. Also worth noting is that MI-only was not tested in the current study given our prior research showing its inferiority to MI+HealthCall in reducing drinking among alcohol dependent PLWH^26^. This study also has several strengths. Our sample included both men and women and members of disadvantaged racial/ethnic groups over-represented in the HIV epidemic. Participant retention was excellent over a lengthy 12-month follow-up. All groups reduced drinking, and CG was well-received by the clinic staff members who administered it. In fact, anecdotally, other clinic staff sought CG training when they heard about the results of the study. Given the recent embrace of telehealth to supplement in-person treatment, HealthCall offers an additional opportunity for technology to extend the impact of in-person or brief online counseling sessions, potentially representing an important tool during the COVID-19 era and beyond.

In sum, among alcohol dependent PLWH, HealthCall paired with MI resulted in significantly less drinking *during* treatment, and HealthCall paired with CG resulted in significantly less drinking *after* treatment. Drinking reduction is an important target for individuals with HIV, as drinking can both contribute to disease processes and interrupt continuity of care. Because HealthCall also targets other important health behaviors, including medication adherence, it offers the potential to make a broader impact on patients’ health. Given the importance of drinking reduction and medication adherence among PLWH, and the low personnel demands of HealthCall, pairing HealthCall with brief evidence-based behavioral interventions in HIV primary care settings merits widespread consideration.

## Data Availability

Participant data is confidential and protected. Please contact with questions.

## References

1. Rehm J, Shield KD. Global Burden of Alcohol Use Disorders and Alcohol Liver Disease. Biomedicines. 2019;7(4).

2. Shield K, Manthey J, Rylett M, et al. National, regional, and global burdens of disease from 2000 to 2016 attributable to alcohol use: a comparative risk assessment study. Lancet Public Health. 2020;5(1):e51–e61.

3. Blanco C, Wall MM, Compton WM, et al. Prevalence and correlates of HIV testing and HIV-positive status in the US: Results from the National Epidemiological Survey on Alcohol and Related Conditions III (NESARC-III). J Psychiatr Res. 2018;105:1-8.

4. Shiau S, Arpadi SM, Yin MT, Martins SS. Patterns of drug use and HIV infection among adults in a nationally representative sample. Addict Behav. 2017;68:39-44.

5. Hartzler B, Dombrowski JC, Crane HM, et al. Prevalence and Predictors of Substance Use Disorders Among HIV Care Enrollees in the United States. AIDS Behav. 2017;21(4):1138–1148.

6. Crane HM, McCaul ME, Chander G, et al. Prevalence and Factors Associated with Hazardous Alcohol Use Among Persons Living with HIV Across the US in the Current Era of Antiretroviral Treatment. AIDS Behav. 2017;21(7):1914–1925.

7. Williams EC, Hahn JA, Saitz R, Bryant K, Lira MC, Samet JH. Alcohol Use and Human Immunodeficiency Virus (HIV) Infection: Current Knowledge, Implications, and Future Directions. Alcohol Clin Exp Res. 2016;40(10):2056–2072.

8. Williams EC, McGinnis KA, Edelman EJ, et al. Level of Alcohol Use Associated with HIV Care Continuum Targets in a National U.S. Sample of Persons Living with HIV Receiving Healthcare. AIDS Behav. 2019;23(1):140–151.

9. Vagenas P, Azar MM, Copenhaver MM, Springer SA, Molina PE, Altice FL. The Impact of Alcohol Use and Related Disorders on the HIV Continuum of Care: a Systematic Review : Alcohol and the HIV Continuum of Care. Curr HIV/AIDS Rep. 2015;12(4):421–436.

10. Korthuis PT, Saha S, Chander G, et al. Substance use and the quality of patient-provider communication in HIV clinics. AIDS Behav. 2011;15(4):832–841.

11. Rehm J, Shield KD, Joharchi N, Shuper PA. Alcohol consumption and the intention to engage in unprotected sex: systematic review and meta-analysis of experimental studies. Addiction. 2012;107(1):51–59.

12. Loutfy MR, Wu W, Letchumanan M, et al. Systematic review of HIV transmission between heterosexual serodiscordant couples where the HIV-positive partner is fully suppressed on antiretroviral therapy. PLoS One. 2013;8(2).

13. Miller WR, Baca C, Compton WM, et al. Addressing substance abuse in health care settings. Alcohol Clin Exp Res. 2006;30(2):292–302.

14. Saitz R. Alcohol screening and brief intervention in primary care: Absence of evidence for efficacy in people with dependence or very heavy drinking. Drug Alcohol Rev. 2010;29(6):631–640.

15. Beyer FR, Campbell F, Bertholet N, et al. The Cochrane 2018 Review on Brief Interventions in Primary Care for Hazardous and Harmful Alcohol Consumption: A Distillation for Clinicians and Policy Makers. Alcohol Alcohol. 2019;54(4):417–427.

16. Hasin DS, Aharonovich E, O’Leary A, et al. Reducing Heavy Drinking in HIV Primary Care: A Randomized Trial of Brief Intervention, with and without Technological Enhancement. Addiction. 2013;108(7):1230–1240.

17. Aharonovich E, Hatzenbuehler ML, Johnston B, et al. A low-cost, sustainable intervention for drinking reduction in the HIV primary care setting. AIDS Care. 2006;18(6):561–568.

18. Madhombiro M, Musekiwa A, January J, Chingono A, Abas M, Seedat S. Psychological interventions for alcohol use disorders in people living with HIV/AIDS: a systematic review. Syst Rev. 2019;8(1):244.

19. Brown JL, DeMartini KS, Sales JM, Swartzendruber AL, DiClemente RJ. Interventions to reduce alcohol use among HIV-infected individuals: a review and critique of the literature. Curr HIV/AIDS Rep. 2013;10(4):356–370.

20. Satre DD, Leibowitz AS, Leyden W, et al. Interventions to Reduce Unhealthy Alcohol Use among Primary Care Patients with HIV: the Health and Motivation Randomized Clinical Trial. J Gen Intern Med. 2019;34(10):2054–2061.

21. Dawson-Rose C, Draughon JE, Cuca Y, et al. Changes in Specific Substance Involvement Scores among SBIRT recipients in an HIV primary care setting. Addict Sci Clin Pract. 2017;12(1):34.

22. Chambers JE, Brooks AC, Medvin R, et al. Examining multi-session brief intervention for substance use in primary care: research methods of a randomized controlled trial. Addict Sci Clin Pract. 2016;11(1):8.

23. Kahler CW, Surace A, Durst A, et al. Telehealth interventions to reduce alcohol use in men with HIV who have sex with men: Protocol for a factorial randomized controlled trial. Contemp Clin Trials Commun. 2019;16:100475.

24. Gilbert P, Ciccarone D, Gansky SA, et al. Interactive “Video Doctor” counseling reduces drug and sexual risk behaviors among HIV-positive patients in diverse outpatient settings. PLoS One. 2008;3(4):e1988.

25. Mgbako O, Miller EH, Santoro AF, et al. COVID-19, Telemedicine, and Patient Empowerment in HIV Care and Research. AIDS Behav. 2020;24(7):1990–1993.

26. Aharonovich E, Greenstein E, O’Leary A, Johnston B, Seol SG, Hasin DS. HealthCall: technology-based extension of motivational interviewing to reduce non-injection drug use in HIV primary care patients - a pilot study. AIDS Care. 2012;24(12):1461–1469.

27. Whitlock EP, Polen MR, Green CA, Orleans T, Klein J, Force USPST. Behavioral counseling interventions in primary care to reduce risky/harmful alcohol use by adults: a summary of the evidence for the U.S. Preventive Services Task Force. Ann Intern Med. 2004;140(7):557–568.

28. Miller WR, Rose GS. Toward a theory of motivational interviewing. Am Psychol. 2009;64(6):527–537.

29. Apodaca TR, Longabaugh R. Mechanisms of change in motivational interviewing: a review and preliminary evaluation of the evidence. Addiction. 2009;104(5):705–715.

30. Hettema J, Steele J, Miller WR. Motivational interviewing. Annu Rev Clin Psychol. 2005;1:91–111.

31. Dunn C, Deroo L, Rivara FP. The use of brief interventions adapted from motivational interviewing across behavioral domains: a systematic review. Addiction. 2001;96(12):1725–1742.

32. Burke BL, Arkowitz H, Menchola M. The efficacy of motivational interviewing: a meta-analysis of controlled clinical trials. J Consult Clin Psychol. 2003;71(5):843–861.

33. Bien TH, Miller WR, Tonigan JS. Brief interventions for alcohol problems: a review. Addiction. 1993;88(3):315–335.

34. Burke BL, Vassilev G, Kantchelov A, Zweben A. Motivational interviewing with couples. In: Miller WR, Rollnick S, eds. Motivational Interviewing: Preparing People for Change. 2nd ed. New York: The Guilford Press; 2002:347–361.

35. Miller WR, Brown J, Simpson TL, et al. What Works? A methodological analysis of the alcohol treatment outcome in literature. In: Hester RK, Miller WR, eds. Handbook of Alcoholism Treatment Approaches: Effective Alternatives. 2nd ed. Boston: Allyn and Bacon; 1995:12–44.

36. Lundahl B, Moleni T, Burke BL, et al. Motivational interviewing in medical care settings: a systematic review and meta-analysis of randomized controlled trials. Patient Educ Couns. 2013;93(2):157–168.

37. Hasin DS, Aharonovich E, Greenstein E. HealthCall for the smartphone: technology enhancement of brief intervention in HIV alcohol dependent patients. Addict Sci Clin Pract. 2014;9:5.

38. Soderlund LL, Madson MB, Rubak S, Nilsen P. A systematic review of motivational interviewing training for general health care practitioners. Patient Educ Couns. 2011;84(1):16–26.

39. Forsberg L, Forsberg LG, Lindqvist H, Helgason AR. Clinician acquisition and retention of Motivational Interviewing skills: a two-and-a-half-year exploratory study. Subst Abuse Treat Prev Policy. 2010;5:8.

40. Pirlott AG, Kisbu-Sakarya Y, Defrancesco CA, Elliot DL, Mackinnon DP. Mechanisms of motivational interviewing in health promotion: a Bayesian mediation analysis. Int J Behav Nutr Phys Act. 2012;9(1):69.

41. Gaume J, Gmel G, Faouzi M, Daeppen JB. Counselor skill influences outcomes of brief motivational interventions. J Subst Abuse Treat. 2009;37(2):151–159.

42. U.S. Department of Health and Human Services, National Institutes of Health, National Institue on Alcohol Abuse and Alcoholism. Helping patients who drink too much: a clinician’s guide. In: Bethesda (MD): National Institue on Alcohol Abuse and Alcoholism; 2007: Available from:http://pubs.niaaa.nih.gov/publications/Practitioner/CliniciansGuide2005/guide.pdf.

43. Willenbring ML, Massey SH, Gardner MB. Helping patients who drink too much: an evidence-based guide for primary care clinicians. Am Fam Physician. 2009;80(1):44–50.

44. Chander G. Addressing alcohol use in HIV-infected persons. Top Antivir Med. 2011;19(4):143–147.

45. National Institute on Alcohol Abuse and Alcoholism. NIAAA Clinician’s Guide Online Training/Video Cases: Helping Patients Who Drink Too Much. http://www.niaaa.nih.gov/publications/clinical-guides-and-manuals/niaaa-clinicians-guide-online-training. Accessed Accessed: July 8, 2013.

46. U.S. Preventive Service Task Force. Screening and Behavioral Counseling Interventions in Primary Care to Reduce Alcohol Misuse: Recommendation Statement. April 2004; Available from: http://www.uspreventiveservicestaskforce.org/3rduspstf/alcohol/alcomisrs.htm.

47. Whitlock EP, Green CA, Polen MR, et al. Behavioral Counseling Interventions in Primary Care to Reduce Risky/Harmful Alcohol Use [Internet]. In: Rockville (MD): Agency for Healthcare Research and Quality (US); 2004: Available from: http://www.ncbi.nlm.nih.gov/books/NBK42870/. Accessed 7/8/2013.

48. Babor TF, McRee BG, Kassebaum PA, Grimaldi PL, Ahmed K, Bray J. Screening, Brief Intervention, and Referral to Treatment (SBIRT): toward a public health approach to the management of substance abuse. Subst Abus. 2007;28(3):7–30.

49. Michie S, Whittington C, Hamoudi Z, Zarnani F, Tober G, West R. Identification of behaviour change techniques to reduce excessive alcohol consumption. Addiction. 2012;107(8):1431–1440.

50. A cross-national trial of brief interventions with heavy drinkers. WHO Brief Intervention Study Group. Am J Public Health. 1996;86(7):948–955.

51. Cuijpers P, Riper H, Lemmers L. The effects on mortality of brief interventions for problem drinking: a meta-analysis. Addiction. 2004;99(7):839–845.

52. Kaner EF, Dickinson HO, Beyer F, et al. The effectiveness of brief alcohol interventions in primary care settings: a systematic review. Drug Alcohol Rev. 2009;28(3):301–323.

53. Bertholet N, Daeppen JB, Wietlisbach V, Fleming M, Burnand B. Reduction of alcohol consumption by brief alcohol intervention in primary care: systematic review and meta-analysis. Arch Intern Med. 2005;165(9):986–995.

54. Hasin DS, Wall M, Witkiewitz K, et al. Change in non-abstinent WHO drinking risk levels and alcohol dependence: a 3 year follow-up study in the US general population. Lancet Psychiatry. 2017;4(6):469–476.

55. Ehrmann J, Urban O, Dvoran P. Alcohol-related liver diseases. Cent Eur J Public Health. 2019;27 Suppl:S10-S14.

56. American Psychiatric Association. Diagnostic and statistical manual of mental disorders (4th ed., text rev.). Washington DC2000.

57. Zhao W, Weng Y. Block urn design - a new randomization algorithm for sequential trials with two or more treatments and balanced or unbalanced allocation. Contemp Clin Trials. 2011;32(6):953–961.

58. Miller W, Rollnick S. Motivational Interviewing: Preparing People for Change, 2nd Edition. The Guilford Press; 2002.

59. Sullivan JT, Sykora K, Schneiderman J, Naranjo CA, Sellers EM. Assessment of alcohol withdrawal: the revised clinical institute withdrawal assessment for alcohol scale (CIWA-Ar). Br J Addict. 1989;84(11):1353–1357.

60. Sobell LC, Sobell MB. Alcohol Consumption Measures. In: Assessing Alcohol Problems: A Guide for Clinicians and Researchers. Washington, D.C.: U.S. Department of Health and Human Services; 1995:55–73.

61. Alcoholism NIoAAa. What Is A Standard Drink? 2017.

62. Delker E, Aharonovich E, Hasin D. Interviewer-administered TLFB vs. self-administered computerized (A-CASI) drug use frequency questions: a comparison in HIV-infected drug users. Drug Alcohol Depend. 2016;161:29–35.

63. Robinson SM, Sobell LC, Sobell MB, Leo GI. Reliability of the Timeline Followback for cocaine, cannabis, and cigarette use. Psychol Addict Behav. 2014;28(1):154–162.

64. Thursz M, Kamath PS, Mathurin P, Szabo G, Shah VH. Alcohol-related liver disease: Areas of consensus, unmet needs and opportunities for further study. J Hepatol. 2019;70(3):521–530.

65. SAS Institute Inc. SAS [computer program]. Version 9.4. In. Cary, NC. 2014.

66. Hasin DS, Aharonovich E, O’Leary A, et al. Reducing heavy drinking in HIV primary care: a randomized trial of brief intervention, with and without technological enhancement. Addiction. 2013;108(7):1230–1240.

67. Hasin D, Aharonovich E, Zingman B, Schlesinger M, Walsh C, Knox J. Smartphone Intervention to Reduce Heavy Drinking in HIV Care: Effect on ART Adherence. Poster presented at: Conference on Retroviruses and Opportunistic Infections (CROI). Presented virtually. March 8–11 2020.

